# Timing of elective tracheotomy and duration of mechanical ventilation amongst patients admitted to intensive care with severe COVID-19: a multicentre prospective cohort study

**DOI:** 10.1101/2021.01.22.21249651

**Authors:** Albert Prats-Uribe, Marc Tobed, José Miguel Villacampa, Adriana Agüero, Clara García-Bastida, José Ignacio Tato, Laura Rodrigáñez, Victoria Duque Holguera, Estefanía Hernández-García, Daniel Poletti, Gabriela Simonetti, Vanessa Villarraga, Carla Meler-Claramonte, Álvaro Sánchez Barrueco, Carlos Chiesa-Estomba, Maria Casasayas, Pablo Parente-Arias, Pedro Castro, Daniel Prieto-Alhambra, Isabel Vilaseca, Francesc Xavier Avilés-Jurado, TraqueoCOVID SEORL Group

## Abstract

**Background:** The COVID-19 pandemic has strained intensive care unit (ICU) resources. Tracheotomy is the most frequent surgery performed on ICU patients and can affect the duration of ICU care. We studied the association between when tracheotomy occurs and weaning from mechanical ventilation, mortality, and intraoperative and postoperative complications.

**Methods:** Multicentre prospective cohort including all COVID-19 patients admitted to ICUs in 36 hospitals in Spain who received invasive mechanical ventilation and tracheotomy between 11 March and 20 July 2020. We used a target emulation trial framework to study the causal effects of early (7 to 10 days post-intubation) versus late (>10 days) tracheotomy on time from tracheotomy to weaning, postoperative mortality, and tracheotomy complications. Cause-specific Cox models were used for the first two outcomes and Poisson regression for the third, all adjusted for potential confounders.

**Findings:** We included 696 patients, of whom 142 (20·4%) received early tracheotomy. Using late tracheotomy as the reference group, multivariable cause-specific analysis showed that early tracheotomy was associated with faster post-tracheotomy weaning (fully adjusted hazard ratio (HR) [95% confidence interval (CI)]: 1·31 [1·02 to 1·81]) without differences in mortality (fully adjusted HR [95% CI]: 0·91 [0·56 to 1·47]) or intraoperative or postoperative complications (adjusted rate ratio [95% CI]: 0·21 [0·03 to 1·57] and 1·49 [0·99 to 2·24], respectively).

**Interpretation:** Early tracheotomy reduced post-tracheotomy weaning time, resulting in fewer mechanical ventilation days and shorter ICU stays, without changing complication or mortality rates. These results support early tracheotomy for COVID-19 patients when clinically indicated.

**Funding:** Supported by the NIHR, FAME, and MRC.

**Research in context:** Evidence before this study
The optimal timing of tracheotomy for critically ill COVID-19 patients remains controversial. Existing guidelines and recommendations are based on limited experiences with SARS-CoV-1 and expert opinions derived from situations that differ from a pandemic outbreak. Most of the available guidance recommends late tracheotomy (>14 days), mainly due to the potential risk of infection for the surgical team and the high patient mortality rate observed early in the first wave of the COVID-19 pandemic. Recent publications have shown that surgical teams can safely perform tracheotomies for COVID-19 patients if they use adequate personal protective equipment. Early tracheotomy seems to reduce the length of invasive mechanical ventilation without increasing complications, which may release crucial intensive care unit (ICU) beds sooner. The current recommendations do not suggest an optimal time for tracheotomy for COVID-19 patients, and no study has provided conclusions based on objective clinical parameters.

Added value of this study
This is the first study aiming to establish the optimal timing for tracheotomy for critically ill COVID-19 patients requiring invasive mechanical ventilation (IMV). The study prospectively recruited a large multicentre cohort of 696 patients under IMV due to COVID-19 and collected data about the severity of respiratory failure, clinical and ventilatory parameters, and whether patients need to be laid flat during their ICU stay (proned). The analysis focused on the duration of IMV, mortality, and complication rates. We used a prospective cohort study design to compare the ‘exposures’ of early (performed at day 7 to 10 after starting IMV) versus late (performed after day 10) tracheotomy and set the treatment decision time on the 7th day after orotracheal intubation.

Implications of all the available evidence
The evidence suggests that tracheotomy within 10 days of starting COVID-19 patients on mechanical ventilation allows these patients to be removed from ventilation and discharged from ICU quicker than later tracheotomy, without added complications or increased mortality. This evidence may help to release ventilators and ICU beds more quickly during the pandemic.

## Background

Tracheotomy is the most common procedure performed for patients in the intensive care unit (ICU), required by 10% to 24% of patients under invasive mechanical ventilation (IMV) for prolonged respiratory support or weaning (1). Although substantial variation in the type and timing of this procedure has been reported (2), some studies have suggested that performing an early tracheotomy may reduce the lengths of IMV and ICU care required (3,4).

About 3% of patients hospitalised with COVID-19 (5,6) suffer from respiratory failure and require IMV. Tracheotomy is therefore the most frequent surgical procedure performed during lockdowns for the SARS-CoV-2 pandemic (7). The main indications for a tracheotomy are long-term intubation, management of secretions, sedation reduction needs, progression to weaning, and prevention of laryngeal oedema. Tracheotomy in these patients minimises the long-term risk of laryngotracheal stenosis and reduces the lengths of mechanical ventilation and ICU stay (8). This last aspect is crucial when ICU space is under strain.

Early in the SARS-CoV-2 pandemic, there were concerns that performing a tracheotomy could put the surgical team at risk of infection. Some centres were unable to perform the procedure as they lacked adequate personal protective equipment (PPE) (9). Published high mortality rates and the difficulties and risks of transferring patients from the ICU to the operation room were also drawbacks (6). Many scientific societies issued guidelines and recommendations so that the procedure could be performed safely for both patient and surgeon (10, 11, 12).Many of the recommendations were based on the experience gained from SARS-CoV-1 and Middle East Respiratory Syndrome and drew on the opinions of expert surgeons and epidemiologists (13). As it was believed that it would be safer to perform the procedure when the patient’s viral load was lower, many guidelines recommended late or very late tracheotomy (14). However, guidance disagreed on which type of tracheotomy was safest (open surgical versus percutaneous) or where it should be performed (15). All guidelines agreed that manoeuvres generating aerosols should be minimised at the time of tracheal entrance to protect surgical teams (9,16,17).

Preliminary data from different centres during the pandemic showed that a tracheotomy could be performed safely, even at the patient’s bedside, if the standard PPE recommendations were followed (18). The complication rates under this scenario seemed to be similar to those reported before the pandemic (18). Some data suggested that early tracheotomy might reduce time to weaning and ICU length of stay (4,18).

We therefore evaluated the effect of disease- and tracheotomy-related variables on the weaning and mortality rates in a large multicentre cohort of COVID-19 ICU patients that required a tracheotomy during IMV. The study was performed in Spain during the first wave of the COVID-19 pandemic and focused on determining the optimal time to perform a tracheotomy for these patients.

## Methods

### Study design and setting

We used a prospective cohort study design. All patients receiving a tracheotomy between 11 March 2020 and 20 July 2020 in 36 hospitals who met the inclusion criteria were included. Fifty patients of the series have been reported elsewhere (18).

A data collection proposal was sent to all members of the Spanish Society of Otorhinolaryngology and Head and Neck Surgery by the senior author (FXA-J). Thirty-six hospitals showed interest. Researchers from each hospital collected the data from ICU admission to weaning/death/end of study and filled in on an MS Excel sheet (MS Excel for mac v16.16.27. Microsoft 2018) sent to each centre at the beginning of study. Once the recruitment period was over, each hospital sent the database anonymously to the coordinator (FXA-J) via a secure server. Treatment, weaning criteria and sedation agents were not standardized among centers.

### Inclusion and exclusion criteria

Patients suffering from respiratory failure caused by SARS CoV-2 infection, confirmed by PCR, requiring IMV and subsequent tracheotomy, performed before 20 July 2020 were included.

Patients with a missing tracheotomy, orotracheal intubation, or outcome date or missing age or sex were excluded. Following our target trial framework, we excluded patients with a tracheotomy performed in the first 7 days after orotracheal intubation.

### Target trial and follow-up

We used a trial emulation framework to minimise confounding and bias. Our exposure was early or late tracheotomy. Our randomisation time (D7) was 7 days after the initiation of IMV (D0), when we expect a decision of whether a patient should have a tracheotomy to be made. All baseline characteristics were considered before or on this date. As all of the study participants received a tracheotomy, an intention-to-treat analysis was impossible and we instead used a per-protocol analysis. We followed-up participants from the day of the tracheotomy (T0) until death, weaning, or the end of 20 July 2020, whichever was sooner. Participants who had not died or weaned by this date were then censored. **Figure 1** describes these timings.

**Figure 1.**
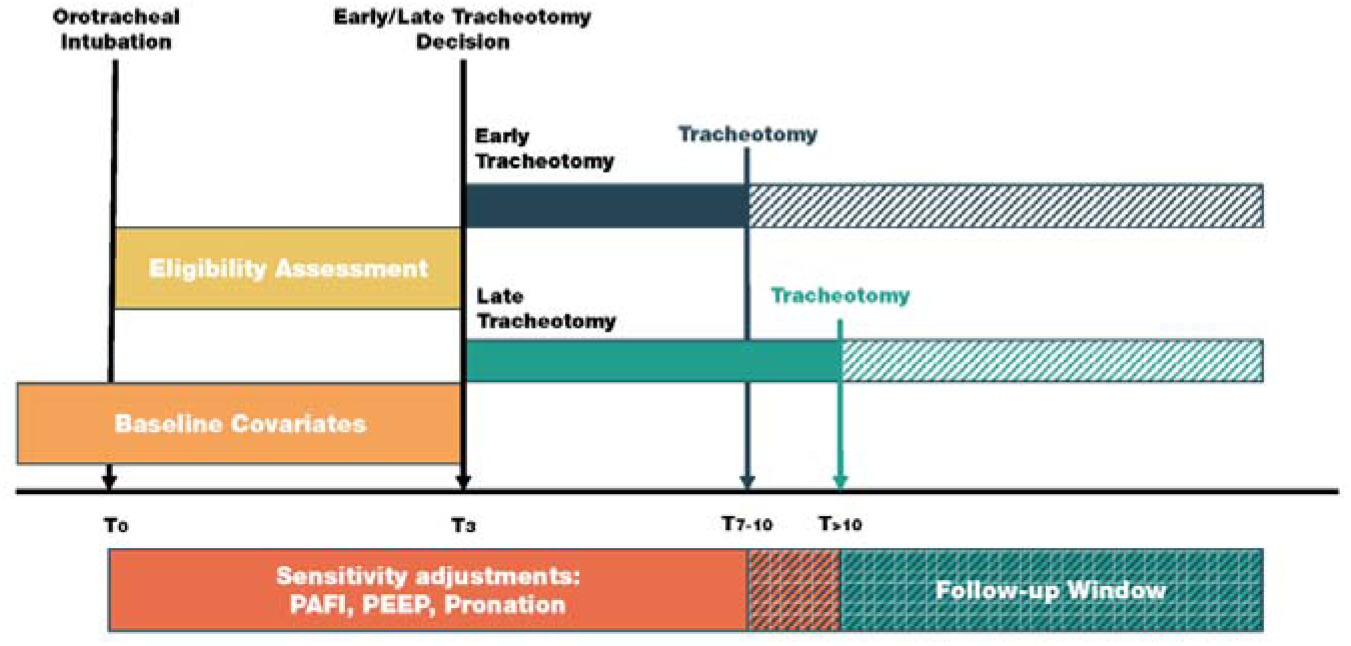
Target trial description. OTI, orotracheal intubation; PAFI, PaO_2_/FiO_2_ ratio; PEEP, positive end espiratory pressure

### Study outcome

The main outcome was time to weaning, defined as days from tracheotomy to weaning from IMV. Secondary outcomes included death, defined as days from tracheotomy to death, and rates of intraoperative bleeding (excessive bleeding that difficult standard tracheotomy or requiring additional haemosthatic measures), postoperative bleeding (bleeding that required revision of stoma) and ventilatory complications (air leak).

### Exposures and measurements

The main exposure variable was early versus late tracheotomy. ‘Early’ was defined as occurring on day 7 to 10 after orotracheal intubation, and ‘late’ as on day 11 or later. Sex and year of birth were acquired at hospital admission. We selected the comorbidities that were most likely to be risk factors for COVID-19 based on previous literature. We included hypertensive disease, immunosuppression, heart failure, autoimmune disease, chronic obstructive pulmonary disease (COPD), pregnancy, diabetes mellitus, neuromuscular disease, and ischaemic heart disease. We registered the start and end days of pronation cycles. We obtained measures of the PaO_2_/FiO_2_ ratio (PAFI) and positive end-expiratory pressure (PEEP) at intubation (D0), 7 days after intubation (decision date, D7), and at tracheotomy (T0). APACHE II and SOFA scores were obtained at ICU admission. We collected the international normalised ratio (INR), use of anticoagulants, use of vasoactive drugs, presence of secretion problems, and indication at surgery. We also collected total lymphocyte and leukocyte count, INR, D-dimer, ferritin, lactate dehydrogenase, and C-reactive protein at admission. These variables are obtained from the electronic medical records, which included analytical parameters, dates of procedures and ventilatory parameters.

### Ethical approval and informed consent

The local ethics committee approved the study protocol and waved informed consent given the observational nature of the study.

### Statistical analysis

We calculated the proportion or mean and standard deviation of each variable for the population as a whole and stratified by exposure and included/excluded status. We computed weekly and total incidence rates (events per 100 person-day) of weaning and death overall and stratified by early versus late tracheotomy. We plotted cumulative incidence curves of weaning and death by exposure. We fitted a multivariable Cox model to estimate cause-specific hazard ratios (csHRs) of weaning and death for early versus late tracheotomy.

We fitted multivariable Poisson models to estimate the relative risk of intraoperative and postoperative bleeding and ventilatory complications. All models were repeat-adjusted for age and sex. All models were further adjusted for age, sex, PAFI, PEEP, and pronation days.

Missing PAFI, PEEP, APACHE II, anticoagulant use, and comorbidity data were imputed using multiple imputation with chained equations. We used predictive mean matching with 5 k nearest neighbours for continuous variables and logistic models dichotomic variables, generating 100 imputed datasets (19). We pooled estimators using Rubin’s rules (20).

We tested for interactions between tracheotomy timing and age, sex, APACHE II, SOFA, PEEP, PAFI, and days of pronation. We compared the participants in the early tracheotomy group who did and did not wean within 14 days of intubation.

We calculated post-hoc power calculations using the Freedman method (21). We performed data management in SPSS 27 (IBM Corp. Released 2020. IBM SPSS Statistics for Windows, Version 27.0. Armonk, NY: IBM Corp). We performed all analyses in STATA version 16.0 (StataCorp. 2019. Stata Statistical Software: Release 16. College Station, TX: StataCorp LLC).

### Study report

We followed the reporting guidelines of the STROBE (Strengthening the Reporting of Observational Studies in Epidemiology) statement for cohort studies. (22).

## Results

Of 794 possible participants, 98 were excluded. Figure 2 shows the flow of patients and numbers included and excluded for each criterion. Supplementary Table 1 compares the characteristics of the included and excluded patients.

**Figure 2.**
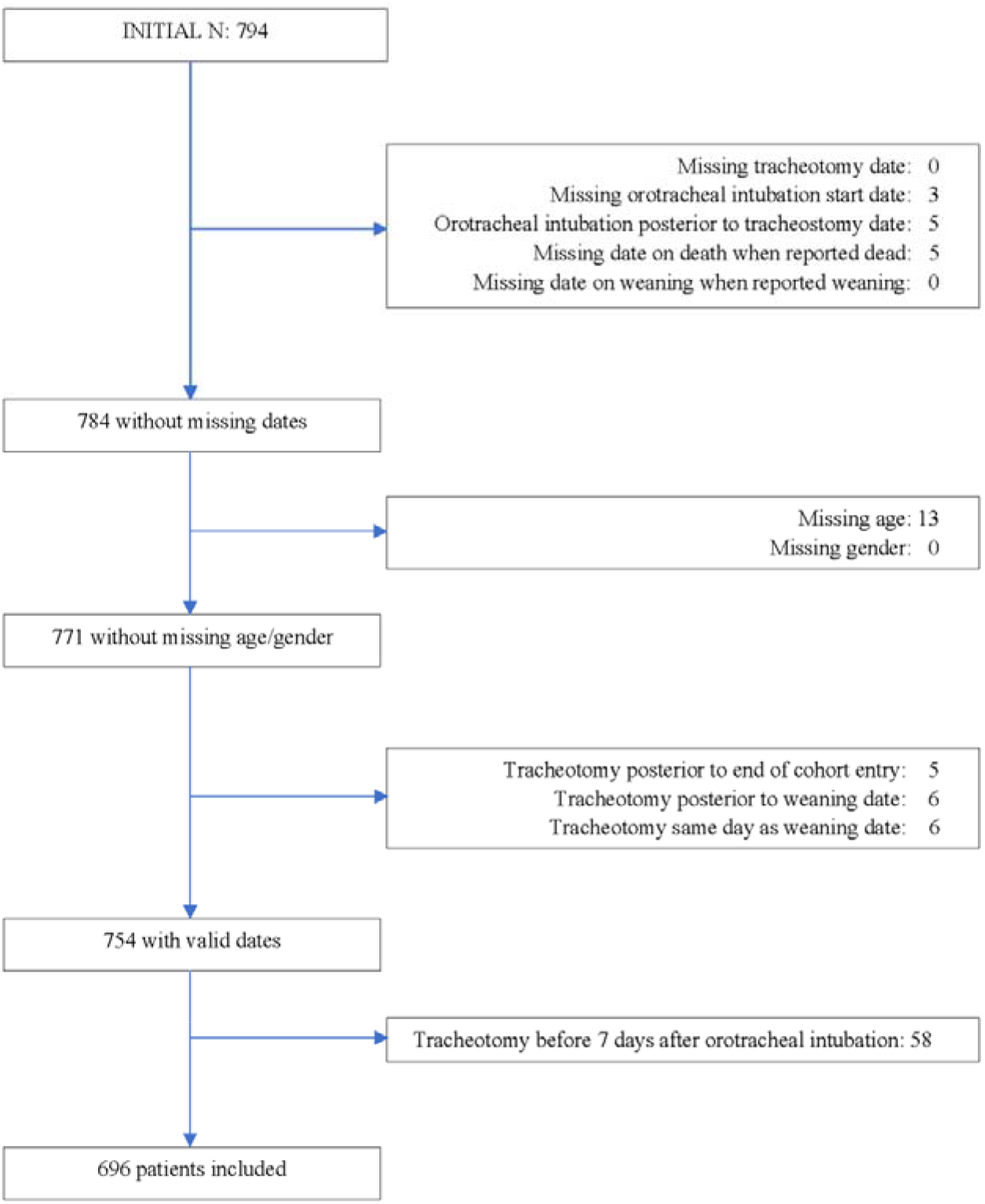
Inclusion and exclusion of study participants.

Table 1 shows the baseline characteristics of the study participants, overall and stratified by tracheotomy timing. Of 696 participants receiving a tracheotomy, 215 (30·9%) were women and 142 (20·4%) received an early tracheotomy. The participants had a mean age of 63 years old. Early and late tracheotomy recipients did not differ on any collected variables except for PAFI at ICU admission, PEEP on tracheotomy day, use of anticoagulant drugs, and days of pronation before tracheotomy.

**Table 1.**
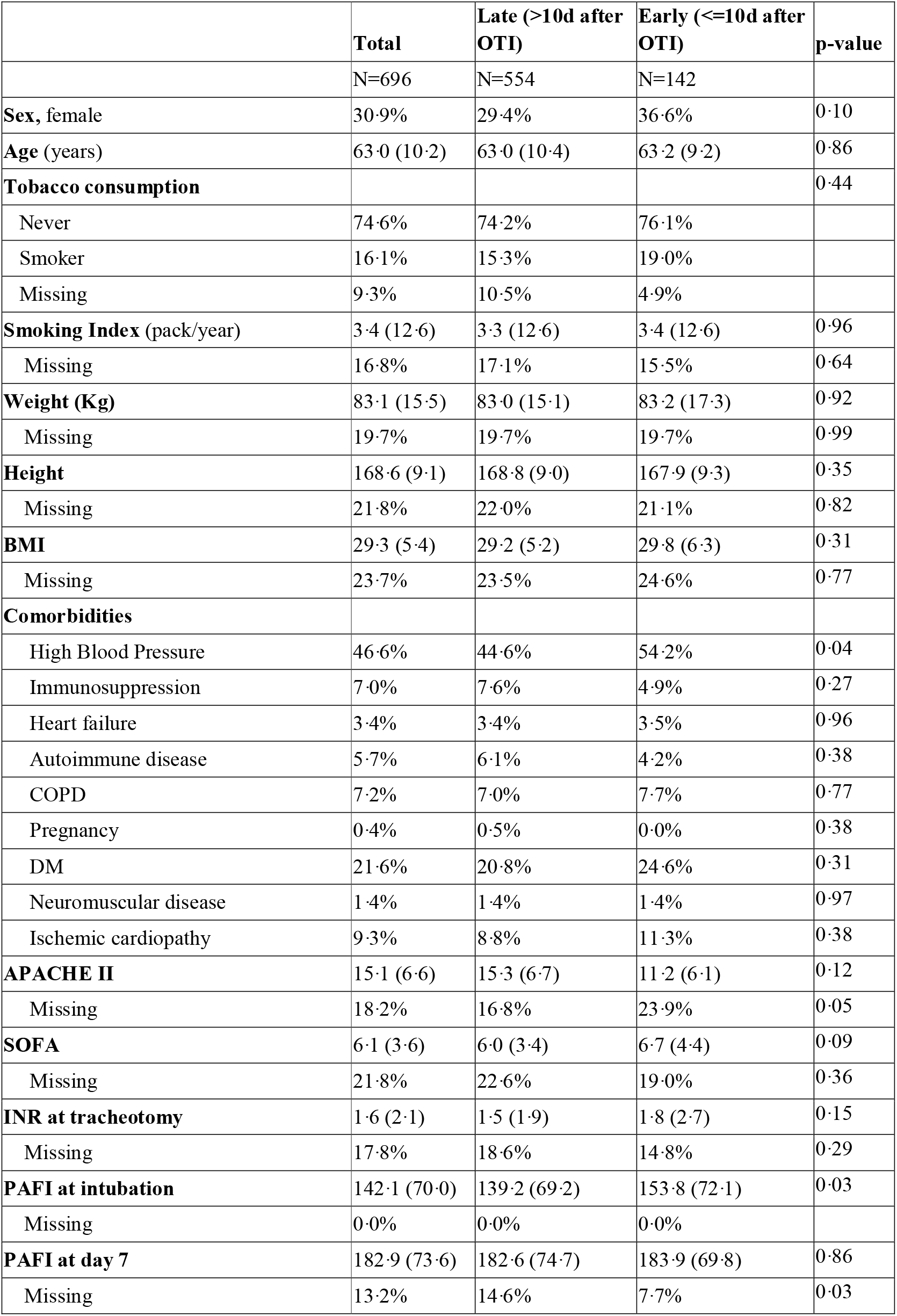
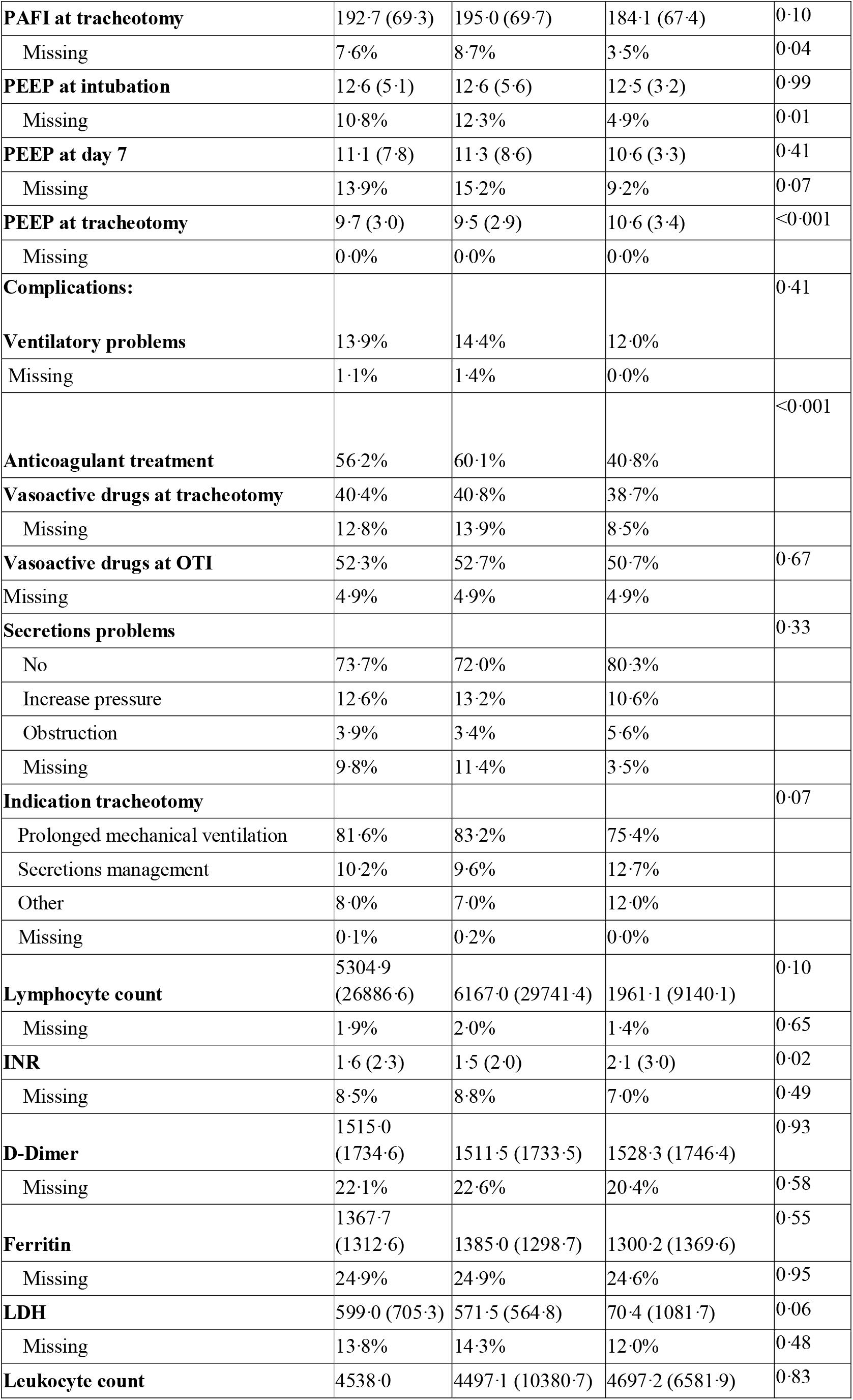
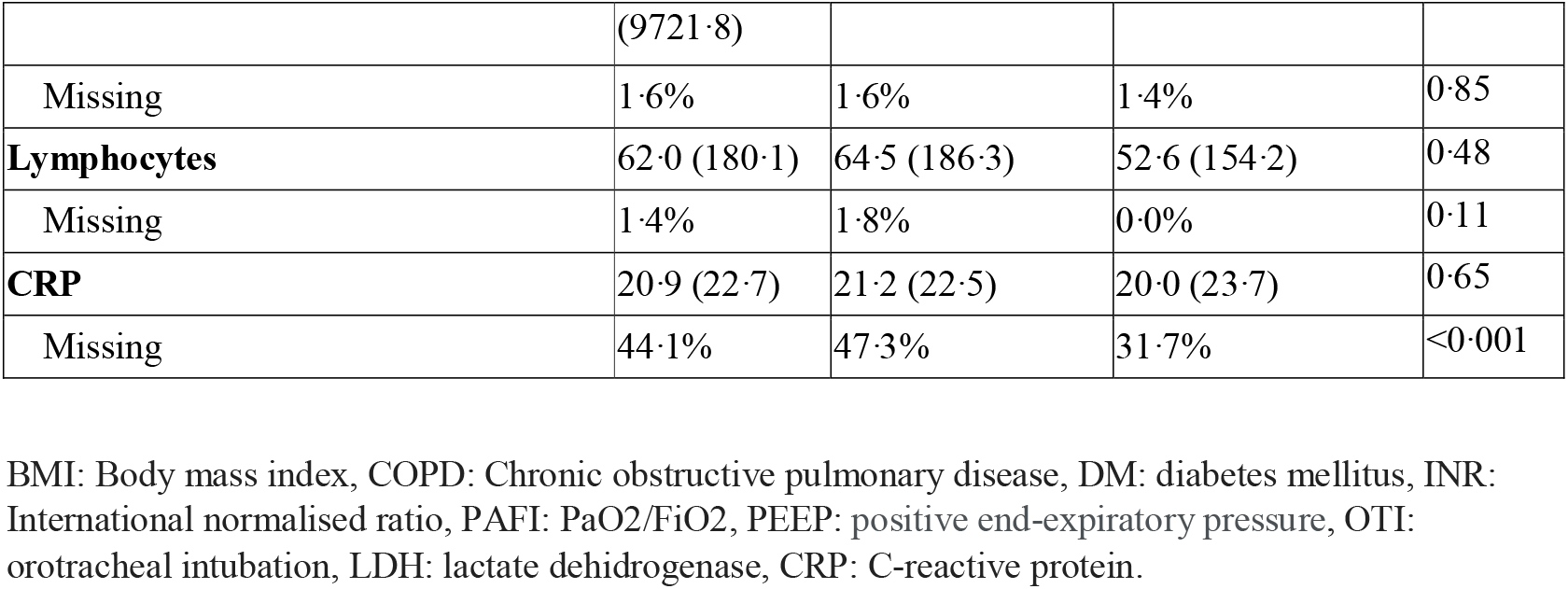
**Baseline characteristics in tracheotomised patients stratified by tracheotomy timing after imputation.**

Supplementary Table 2 compares the frequency of pronation, days of pronation, and whether pronation finished after or before tracheotomy in the two groups. Participants with late tracheotomies had more total days of pronation (9·5 days for late versus 6·8 for early) and more often had their last pronation cycle before tracheotomy (50% for late and 33% for early). The proportion of participants pronated in the first 7 days after orotracheal intubation and how many days these participants were pronated were similar in the late and early tracheotomy groups.

The early tracheotomy group weaned more quickly than the late group (Table 2). The median follow-up time from tracheotomy to weaning or death was 13 days for late tracheotomy and 12 days for early tracheotomy. Among those who were successfully weaned, participants with an early tracheotomy presented 11 days of weaning since tracheotomy and 19 days since orotracheal intubation, whereas late tracheotomy group presented 12 and 29 days, respectively.

**Table 2.** **Days to weaning by tracheotomy timing, since orotracheal intubation and since tracheotomy.**

|  | Early Tracheotomy |  |  | Late Tracheotomy |  |  |
| --- | --- | --- | --- | --- | --- | --- |
| Days to weaning or death or censoring | Median | p25 | p75 | Median | p25 | p75 |
| *Since Tracheotomy (T0)* | 12 | 6 | 22 | 13 | 7 | 26 |
| *Since Decision (D7)* | 13 | 8 | 24 | 23 | 16 | 37 |
| *Since Orotracheal Intubation (D0)* | 20 | 15 | 31 | 30 | 23 | 44 |
| **Days to weaning (those who wean)** |  |  |  |  |  |  |
| *Since Tracheotomy (T0)* | 11 | 6 | 17 | 12 | 7 | 21 |
| *Since Decision (D7)* | 12 | 8 | 20 | 22 | 16 | 33 |
| *Since Orotracheal Intubation (D0)* | 19 | 15 | 27 | 29 | 23 | 40 |
| **Days to death (those who die)** |  |  |  |  |  |  |
| *Since Tracheotomy (T0)* | 13 | 7 | 23 | 14 | 7 | 25 |
| *Since Decision (D7)* | 16 | 9 | 26 | 24 | 17 | 35 |
| *Since Orotracheal Intubation (D0)* | 23 | 16 | 33 | 31 | 24 | 42 |

T0: , D7: , D0: .

The late tracheotomy group had a lower rate of successful weaning: 360 of the 554 participants in the late tracheotomy group were weaned before the end of follow-up (3.1 patients weaning per 100 patient-days [2·8 to 3·4]), whereas 102 out of 142 were weaned in the early tracheotomy group (3·9 patients weaning per 100 patient-days [3·1 to 4·7]). The early tracheotomy group therefore had a higher probability of weaning post-tracheotomy (Table 3).

**Table 3.** **Associations of tracheotomy timing with time to weaning and time to death.**

|  | Weaning |  |  | Death |  |  |
| --- | --- | --- | --- | --- | --- | --- |
| Early vs Late Tracheotomy | csHR | 95% CI |  | csHR | 95% CI |  |
| *Cox* | 1·25 | (1·00 | 1·56) | 0·85 | (0·60 | 1·21) |
| *Cox Age and sex Adjusted* | 1·25 | (1·00 | 1·56) | 0·87 | (0·61 | 1·25) |
| *Cox Fully Adjusted* | 1·31 | (1·02 | 1·81) | 0·91 | (0·56 | 1·47) |

csHR: cause-specific hazard ratio, CI: confidence interval. The fully adjusted model included PAFI (PaO<sub>2</sub>/FiO<sub>2</sub>), PEEP (positive end-expiratory pressure), SOFA (Sequential Organ Failure Assessment
Score) APACHE II (Acute Physiology And Chronic Health Evaluation II Score) and pronation days as covariates.□

Both groups had a mortality rate of 0·32 deaths per 100 patient-days (0·23 to 0·44), with 170/554 participant deaths in the late group and 38/142 in the early group (0·32 deaths per 100 patient-days [0·23 to 0·44]), giving an unadjusted csHR of 0·85 (95% CI: 0·60 to 1·21) and a fully adjusted csHR of 0·91 (0·56 to 1·47). Figure 3 shows cumulative incidence function plots for weaning and death and Table 3 reports csHRs for weaning and death in unadjusted, age- and sex-adjusted, and fully adjusted multivariable models. Early tracheotomy was associated with a risk ratio for intraoperative complications of 0·57 (95% CI: 0·24 to 1·34) in the crude analyses and fully adjusted of 0·21 (0·03 to 1·57) in the adjusted analyses. We did not observe any differences in the risk of any studied intraoperative or postoperative complications between the two groups (Table 4). Participants with early tracheotomy who were weaned in less than 14 days after orotracheal intubation were younger (63·8 versus 60·2 years) and had a higher PAFI at the time of tracheotomy (216·6 versus 177·6) than those who took more than 2 weeks to be weaned (Supplementary Table 4).

**Table 4.** **Associations of tracheotomy timing with incidence of intraoperative and postoperative complications.**

|  | N |  | Crude |  |  | Fully Adjusted |  |  |
| --- | --- | --- | --- | --- | --- | --- | --- | --- |
| **Early vs Late Tracheotomy** | **Early** | **Late** | **RR** | **95% CI** |  | **RR** | **95% CI** |  |
| *Intraoperative* | 6 | 41 | 0.57 | (0.24 | 1.34) | 0.21 | (0.03 | 1.57) |
| *Bleeding* | 4 | 20 | 0.78 | (0.27 | 2.28) | 0.57 | (0.07 | 4.77) |
| *Ventilatory problems* | 4 | 22 | 0.71 | (0.24 | 2.05) | 0.32 | (0.04 | 2.51) |
| *Postoperative* | 41 | 136 | 1.17 | (0.83 | 1.66) | 1.49 | (0.99 | 2.24) |
| *Bleeding* | 33 | 102 | 1.26 | (0.85 | 1.87) | 1.77 | (1.11 | 2.83) |
| *Ventilatory problems* | 8 | 34 | 0.92 | (0.42 | 1.98) | 0.91 | (0.39 | 2.15) |

RR: relative risk, CI: confidence interval. The fully adjusted model included PAFI (PaO<sub>2</sub>/FiO<sub>2</sub>), PEEP (positive end-expiratory pressure), SOFA (Sequential Organ Failure Assessment Score), APACHE II (Acute Physiology And Chronic Health Evaluation II Score), and pronation days as covariates.□

**Figure 3.**
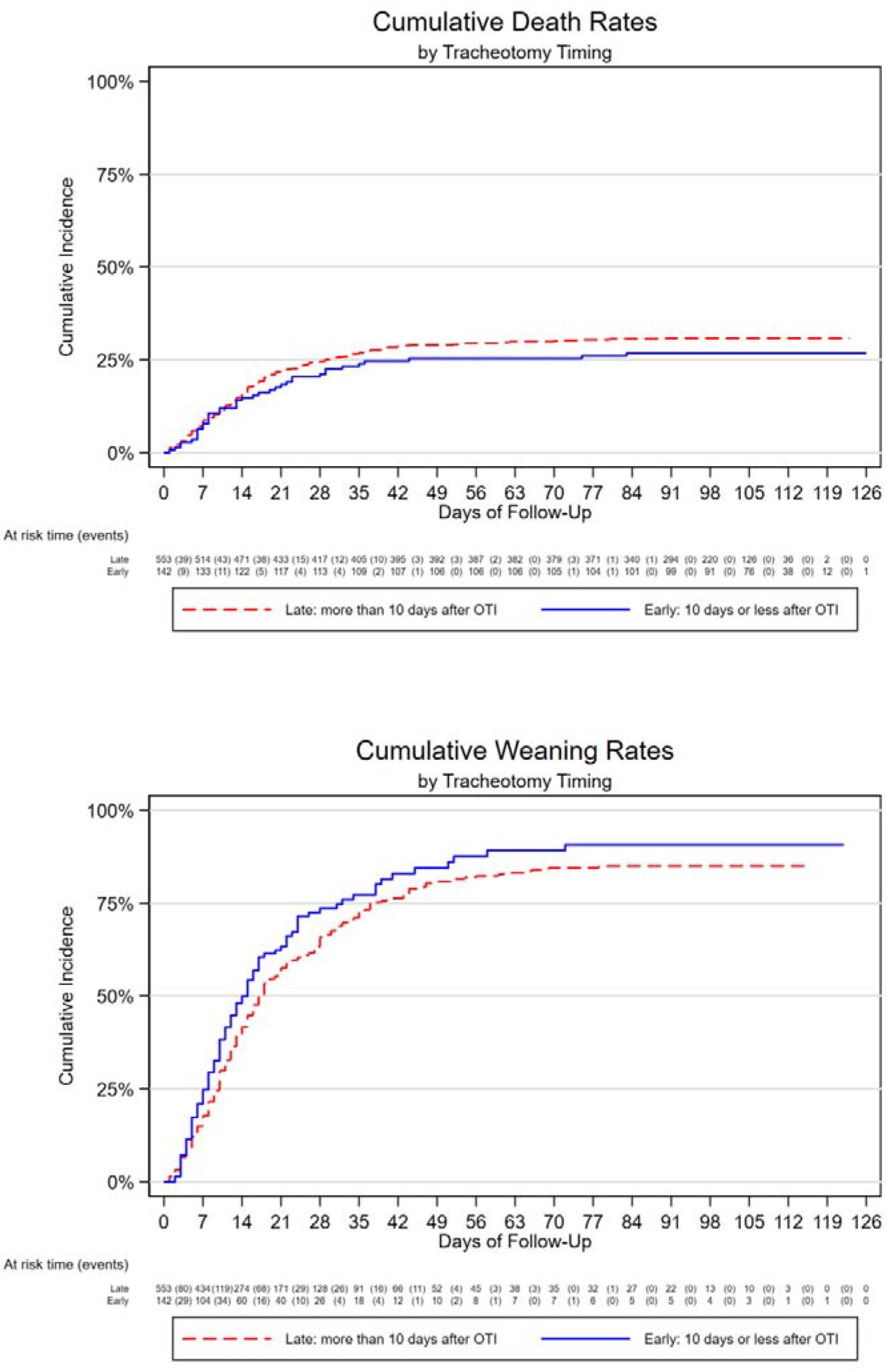
Cumulative incidence of weaning and death by tracheotomy timing.

Using an alpha risk of 5%, the survival seen in each group and the sample size of our cohort, we estimated a statistical power of 79.6%.

## Discussion

To the best of our knowledge, this is the largest study evaluating the optimal timing of tracheotomy in a multicentre prospective cohort of ICU-admitted patients with COVID-19 who required IMV. Our results suggest that early tracheotomy leads to faster weaning without increased complications or mortality. When clinically appropriate, the early tracheotomy strategy might therefore be preferable, as it can release ICU space. The optimal timing of tracheotomy may be influenced by many factors, such as the initial cause of IMV, the severity of the disease, neurological status, complications, and the possibility of recovery. In the COVID-19 scenario, the initial cause of IMV is always severe respiratory failure, which may have high pronation requirements and be complicated by systemic failure, thrombosis, or other COVID-19-related conditions. Many consensus documents were published in 2020 on best practices for tracheotomy in critically ill COVID-19 patients, generally aiming to prevent surgeon infection. Most recommended delayed tracheotomy (14), while others focused on the type of tracheotomy (15), the necessary protective equipment (16,17), or where best to perform a tracheotomy (23). Almost no existing guidance evaluated which clinical parameters influence weaning outcomes, total days of IMV, or mortality.

Bier-Laning and colleagues analysed the tracheotomy protocols and practices put in place by 29 institutions around the world in response to the COVID-19 pandemic. They found insufficient evidence for recommending a specific timing for tracheotomy in COVID-19-related respiratory failure (24). Tornari and colleagues conducted an observational cohort study to understand the factors that influenced the trajectory from tracheotomy to decannulation to facilitate ICU capacity planning and improve outcomes. Higher FiO_2_ at tracheotomy time and higher pre-tracheotomy peak cough flow were associated with longer delays in decannulation of COVID-19 tracheotomy patients (25).

Recent publications have suggested that earlier tracheotomy might facilitate the weaning process and reduce the length of mechanical ventilation required. Avilés-Jurado and colleagues (18) evaluated 50 consecutive patients that required tracheotomy in the first wave of the pandemic in Spain in one single centre. Early tracheotomy reduced the duration of IMV by reducing the days between orotracheal intubation and the tracheotomy procedure, releasing ICU beds for other patients. Our multicentre study of 696 tracheotomised patients from the first wave of COVID-19 confirms their preliminary findings. We found a 31% (2% to 81%) reduction in the time from orotracheal intubation to weaning when tracheotomy was performed early (fully adjusted HR [95% CI]: 1·31 [1·02 to 1·81]), without increasing complications or mortality rates.

The ideal timing for a tracheotomy in patients receiving IMV also remains controversial in other disease scenarios, despite decades of experience of using this technique. Two large randomised prospective studies addressing this topic have been published so far. Terragni and colleagues, randomised 419 patients to early (6 to 9 days of intubation) or late (13 to 15 days of intubation) tracheotomy (26). They found no differences in complications, pneumonia associated with mechanical ventilation or mortality at 28 days of intubation. Young and colleagues randomised 909 patients to early (within 10 days of intubation) or late tracheotomy (after 10 days of intubation) and also found no difference in mortality (27). Neither of these randomised controlled trials examined the days of mechanical ventilation as an outcome. Our results agree with the trials’ finding of no difference in mortality between early and late tracheotomy, but we also found a 31% reduction in IMV duration in the early tracheotomy group.

A Cochrane review published in 2015 found that patients who underwent early (≤10 days) tracheotomy had a higher probability of discharge from the ICU at day 28 than those who underwent later tracheotomy (3). A recent meta-analysis found that early tracheotomy was associated with shorter mechanical ventilation and hospital stays, without differences in mortality (26). Our findings agree with these synthesised results. Reducing IMV and ICU stays is extremely important given the current shortage of ICU beds.

In daily practice, the decision about when to perform a tracheotomy is based on clinical and ventilatory criteria, previous institutional experience, and staff availability to do the procedure. To overcome the lack of a prospective randomised trial, we emulated a trial framework randomising at day 7, which is approximately when the necessity of doing a tracheotomy arises. We included demographic and objective parameters of severity respiratory failure in the analysis. We also included parameters known to influence the intensivist decision such as pronation requirements, PAFI, PEEP levels, age, comorbidities, complications, difficulties in secretions management, and poor prognosis. In our study, treatment, sedation agents and weaning criteria were not standardised among centres. Although it may be seen as a drawback, it represents real practice in the present pandemic scenario, and may give external validity to the cohort. McGrath and colleagues (14) published a consensus document suggesting that tracheotomy be delayed until at least day 10 of mechanical ventilation in COVID-19 patients and only be considered if patients showed signs of clinical improvement. They also advised against tracheotomy when patients needed high fractions of inspired oxygen (FiO_2_), the prone position, or high ventilator requirements. Our participants in the early and late tracheotomy groups had similar characteristics at admission. Although the early group had higher PEEP at tracheotomy day (10·6 versus 9·5, p<0.001), this difference has limited clinical value.

The prone position is often used to improve the ventilation/perfusion quotient in patients with COVID-19. The presence of a tracheotomy can make pronation manoeuvres more difficult, and cannula displacement may also be favoured. Some guidelines suggest tracheotomy be delayed if prone manoeuvres are still required or even discourage tracheotomy altogether (9,14). However, pronation is not a formal contraindication for early tracheotomy. As expected, our late tracheotomy group had greater pronation requirements than our early group. Nonetheless, 63·4% of the early tracheotomy patients were proned at some point, mostly (60·6%) before tracheotomy. Almost 25% continued pronation after tracheotomy without major incidences. Our results therefore suggest that pronation should not rule out early tracheotomy and that patients should be considered on a case-by-case basis.

We found that the shorter IMV duration seen in the early tracheotomy group was independent of ventilatory parameters at admission or tracheotomy. Previous reports based on smaller series have described similar, albeit not statistically significant, findings (18).

One of the criticisms of doing an early tracheotomy is that we may perform more procedures than is strictly necessary. To test this hypothesis, we compared our early tracheotomy participants who were weaned within 2 weeks of intubation (early) with those who were weaned after 2 weeks. Participants who weaned earlier were younger and had higher PAFI at tracheotomy, suggesting that the tracheotomy decision could be delayed in younger patients with clear signs of improvement. One can argue that perhaps some of these patients could have been extubated skipping a tracheotomy, or at least a trial of extubation being performed first. However, in the early days of the pandemic, lack of knowledge about the behaviour of the disease, the risk of infection of staff and the possibility of reintubation, led to a conservative approach to risky manoeuvres.

Another criticism of early tracheotomy is the higher risk of complications in critically ill patients. The high mortality rate initially reported for COVID-19 patients favoured avoiding invasive manoeuvres, including surgery. Many of these patients also required intensive pronation, and some received anticoagulant drugs due to concomitant thrombosis, which may increase tracheotomy complications. Relevant postoperative complications after tracheotomy in non-COVID-19 patients range between 5·6% and 27·2% (28–30). In contrast, Botti and colleagues found that up to 55·3% of tracheotomised COVID-19 patients presented postoperative complications within the first 30 days, with no differences between percutaneous and open procedures (15). The main complications were local infection (36%), haemorrhage (19%), and subcutaneous emphysema (8·5%). Other studies have shown that minor bleeding from the stoma, usually managed with local measures, is the most common complication in tracheotomised COVID-19 patients and that few patients need revision surgery. We found a complication rate of 4·2% for early tracheotomy and 11·2% for late tracheotomy across our 696 participants from different institutions, tracheotomy types, and timelines. This difference was not statistically significant.

Our study has limitations as well as strengths. The study design and inclusion criteria prevented us from analysing the causal effects of tracheotomy timing on total days of IMV. Although the observed differences in total IMV duration post-tracheotomy described here are attributable to the tracheotomy timing, the observed differences in total duration of IMV were artificially inflated by immortal time bias (31) and should not be interpreted as causal estimates. A randomised controlled trial with an intention-to-treat analysis would be preferable to establish the effect of early tracheotomy on the total duration of IMV for patients admitted to ICU with COVID-19, although it is unlikely that it can be performed in the pandemic scenario.

As we used observational data, one can argue that residual confounding could be at least partially responsible for the observed findings. However, although the groups were not totally comparable, we did not find any evidence of confounding by indication, with baseline characteristics well balanced between the early and late tracheotomy groups. Moreover, multivariable adjustment including APACHE II, SOFA, PAFI, PEEP and pronation did not attenuate the observed effects, with a good statistical power.

Among the strengths of the present study are the large number of participants and the prospective cohort design. We designed the analyses following a trial emulation framework (32) with randomisation at day 7 of intubation for robustness. Using multiple centres, each with their own local protocols, no standardized weaning criteria or sedation agents, and variations in the type of tracheotomy, we ensured that this study has external validity. In addition, by collecting multiple objective clinical and ventilatory variables, we were able to gather wide, robust prognosis information. These aspects could have influenced the observed weaning and mortality rates. However, we expect any potential differences to be hospital-specific and therefore uninformative.

In conclusion, our prospective cohort study suggests that early tracheotomy, when appropriate, may provide quicker weaning and ICU discharge for COVID-19 patients without added complications or increased mortality. These findings may help to release ICU beds, which is particularly necessary during the pandemic outbreak.

## Data Availability

The data in the study are the responsibility of the Research Commission of the Spanish Society of ENT

## Acknowledgements

The authors acknowledge English language editing by Dr Jennifer A de Beyer of the Centre for Statistics in Medicine, University of Oxford. We thank the Spanish Society of Otorhinolaryngology-Head Neck Surgery (SEORL-CCC) for facilitating communication between centres.

## Contributorship statement

All authors contributed to the design of the study, interpretation of the results, and manuscript review.

Albert Prats-Uribe, Daniel Prieto-Alhambra, and F. Xavier Avilés-Jurado had access to the data, performed the statistical analysis, and acted as guarantor.

Albert Prats-Uribe wrote the first draft of the manuscript.

F. Xavier Avilés-Jurado and Isabel Vilaseca are the senior authors. The corresponding author attests that all listed authors meet authorship criteria and that no others meeting the criteria have been omitted.

## Transparency declaration

The lead author affirms that the manuscript is an honest, accurate, and transparent account of the study being reported; that no important aspects of the study have been omitted; and that any discrepancies from the study as planned have been explained.

## Data sharing statement

Only necessary for trials

---

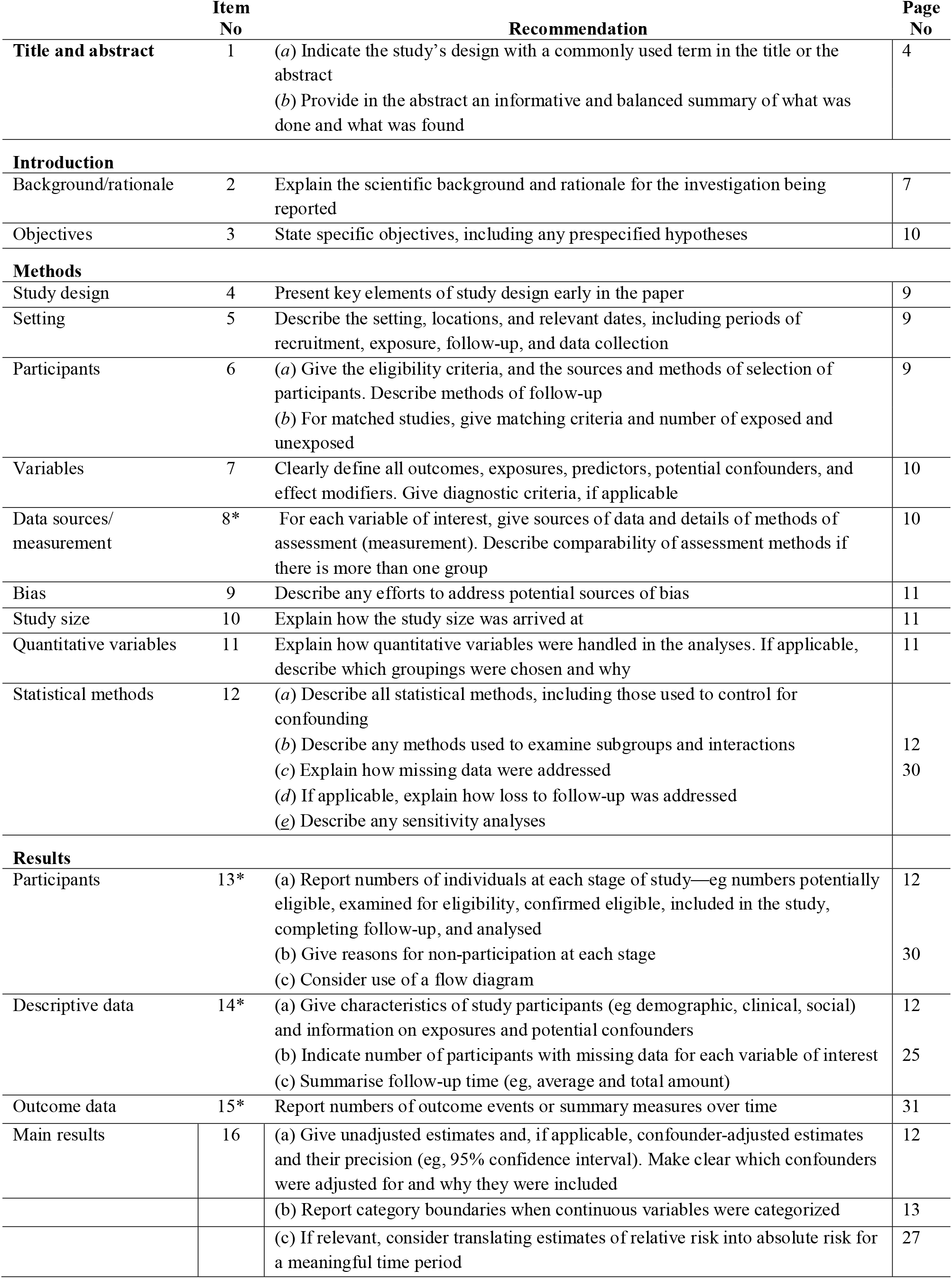
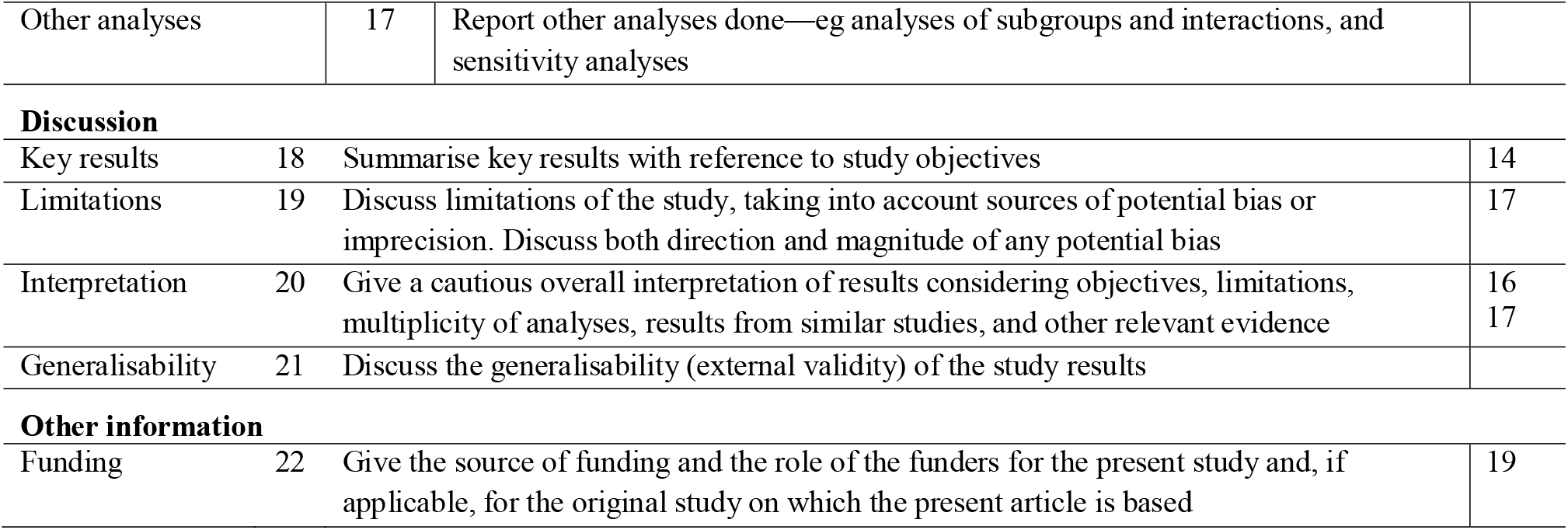

|  | Item No | Recommendation | Page No |
| --- | --- | --- | --- |
| **Title and abstract** | 1 | (a) Indicate the study's design with a commonly used term in the title or the abstract<br>(b) Provide in the abstract an informative and balanced summary of what was done and what was found | 4 |
| **Introduction** |  |  |  |
| Background/rationale | 2 | Explain the scientific background and rationale for the investigation being reported | 7 |
| Objectives | 3 | State specific objectives, including any prespecified hypotheses | 10 |
| **Methods** |  |  |  |
| Study design | 4 | Present key elements of study design early in the paper | 9 |
| Setting | 5 | Describe the setting, locations, and relevant dates, including periods of recruitment, exposure, follow-up, and data collection | 9 |
| Participants | 6 | (a) Give the eligibility criteria, and the sources and methods of selection of participants. Describe methods of follow-up<br>(b) For matched studies, give matching criteria and number of exposed and unexposed | 9 |
| Variables | 7 | Clearly define all outcomes, exposures, predictors, potential confounders, and effect modifiers. Give diagnostic criteria, if applicable | 10 |
| Data sources/<br>measurement | 8* | For each variable of interest, give sources of data and details of methods of assessment (measurement). Describe comparability of assessment methods if there is more than one group | 10 |
| Bias | 9 | Describe any efforts to address potential sources of bias | 11 |
| Study size | 10 | Explain how the study size was arrived at | 11 |
| Quantitative variables | 11 | Explain how quantitative variables were handled in the analyses. If applicable, describe which groupings were chosen and why | 11 |
| Statistical methods | 12 | (a) Describe all statistical methods, including those used to control for confounding<br>(b) Describe any methods used to examine subgroups and interactions<br>(c) Explain how missing data were addressed<br>(d) If applicable, explain how loss to follow-up was addressed<br>(e) Describe any sensitivity analyses | 12<br>30 |
| **Results** |  |  |  |
| Participants | 13* | (a) Report numbers of individuals at each stage of study—eg numbers potentially eligible, examined for eligibility, confirmed eligible, included in the study, completing follow-up, and analysed<br>(b) Give reasons for non-participation at each stage<br>(c) Consider use of a flow diagram | 12<br>30 |
| Descriptive data | 14* | (a) Give characteristics of study participants (eg demographic, clinical, social) and information on exposures and potential confounders<br>(b) Indicate number of participants with missing data for each variable of interest<br>(c) Summarise follow-up time (eg, average and total amount) | 12<br>25 |
| Outcome data | 15* | Report numbers of outcome events or summary measures over time | 31 |
| Main results | 16 | (a) Give unadjusted estimates and, if applicable, confounder-adjusted estimates and their precision (eg, 95% confidence interval). Make clear which confounders were adjusted for and why they were included | 12 |
|  |  | (b) Report category boundaries when continuous variables were categorized | 13 |
|  |  | (c) If relevant, consider translating estimates of relative risk into absolute risk for a meaningful time period | 27 |
| Other analyses | 17 | Report other analyses done—eg analyses of subgroups and interactions, and sensitivity analyses |  |
| **Discussion** |  |  |  |
| Key results | 18 | Summarise key results with reference to study objectives | 14 |
| Limitations | 19 | Discuss limitations of the study, taking into account sources of potential bias or imprecision. Discuss both direction and magnitude of any potential bias | 17 |
| Interpretation | 20 | Give a cautious overall interpretation of results considering objectives, limitations, multiplicity of analyses, results from similar studies, and other relevant evidence | 16<br>17 |
| Generalisability | 21 | Discuss the generalisability (external validity) of the study results |  |
| **Other information** |  |  |  |
| Funding | 22 | Give the source of funding and the role of the funders for the present study and, if applicable, for the original study on which the present article is based | 19 |

## Supplementary Material

**Table S1.**
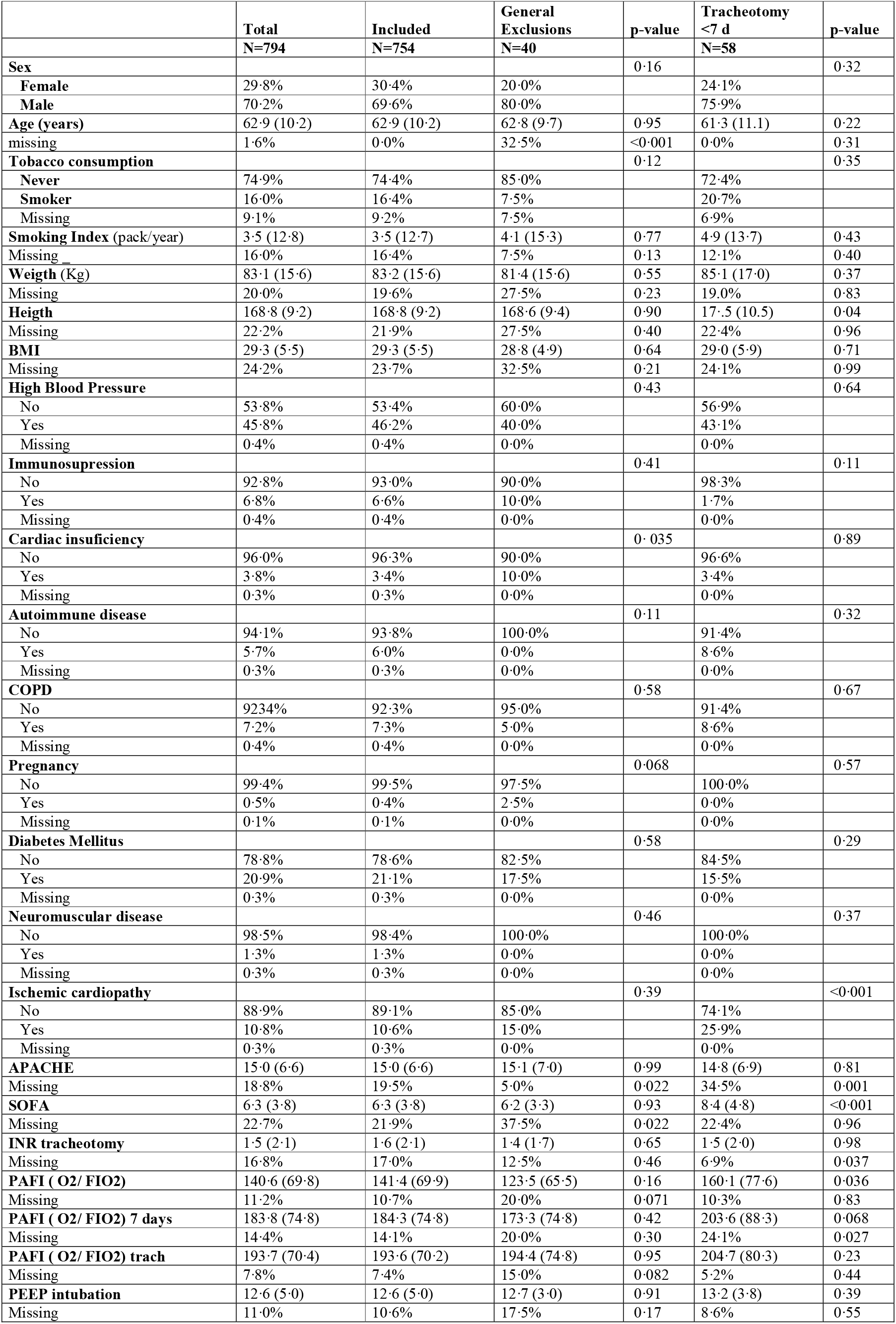
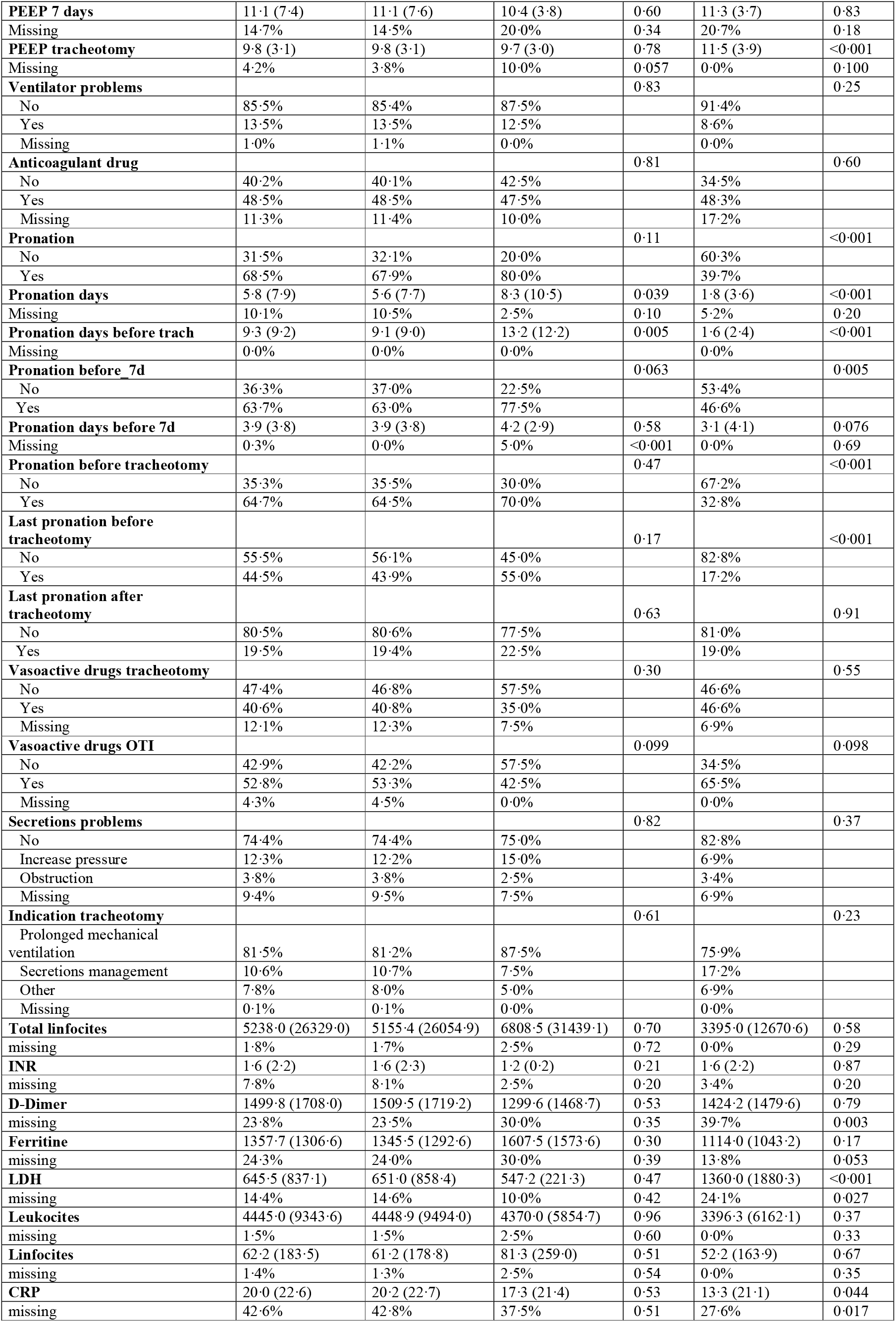
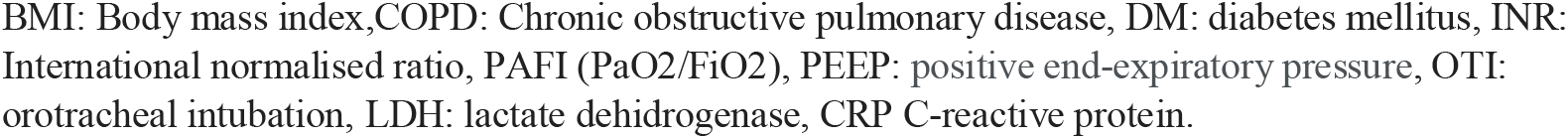
Baseline characteristics in tracheotomised patients stratified by exclusion.

**Table S2.**
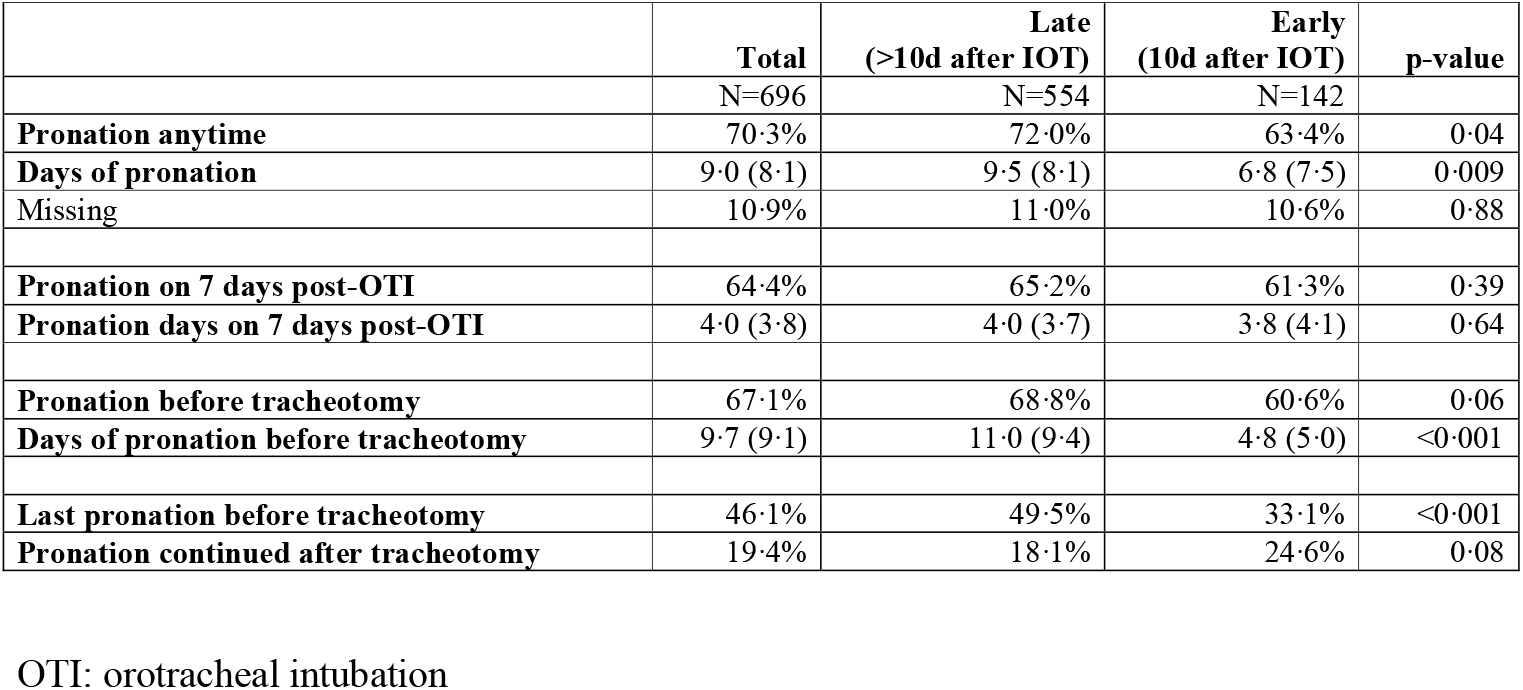
**Pronation and pronation days before tracheotomy in tracheotomised patients stratified by early or late weaning.**

**Figure S3.**
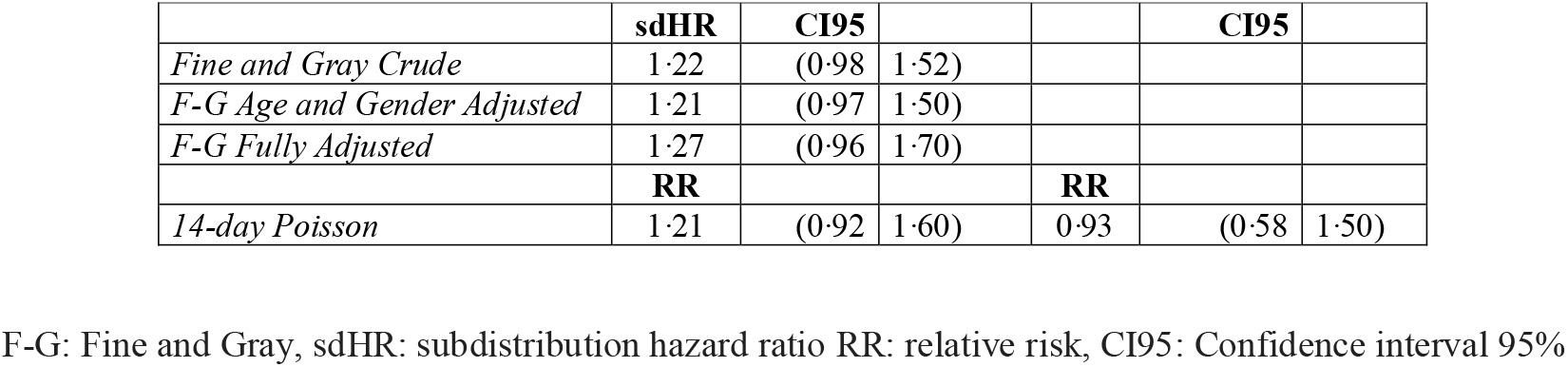
Additional Analyses of time to weaning: Fine and Gray competing risks model and 14-day Poisson.

**Figure S4.**
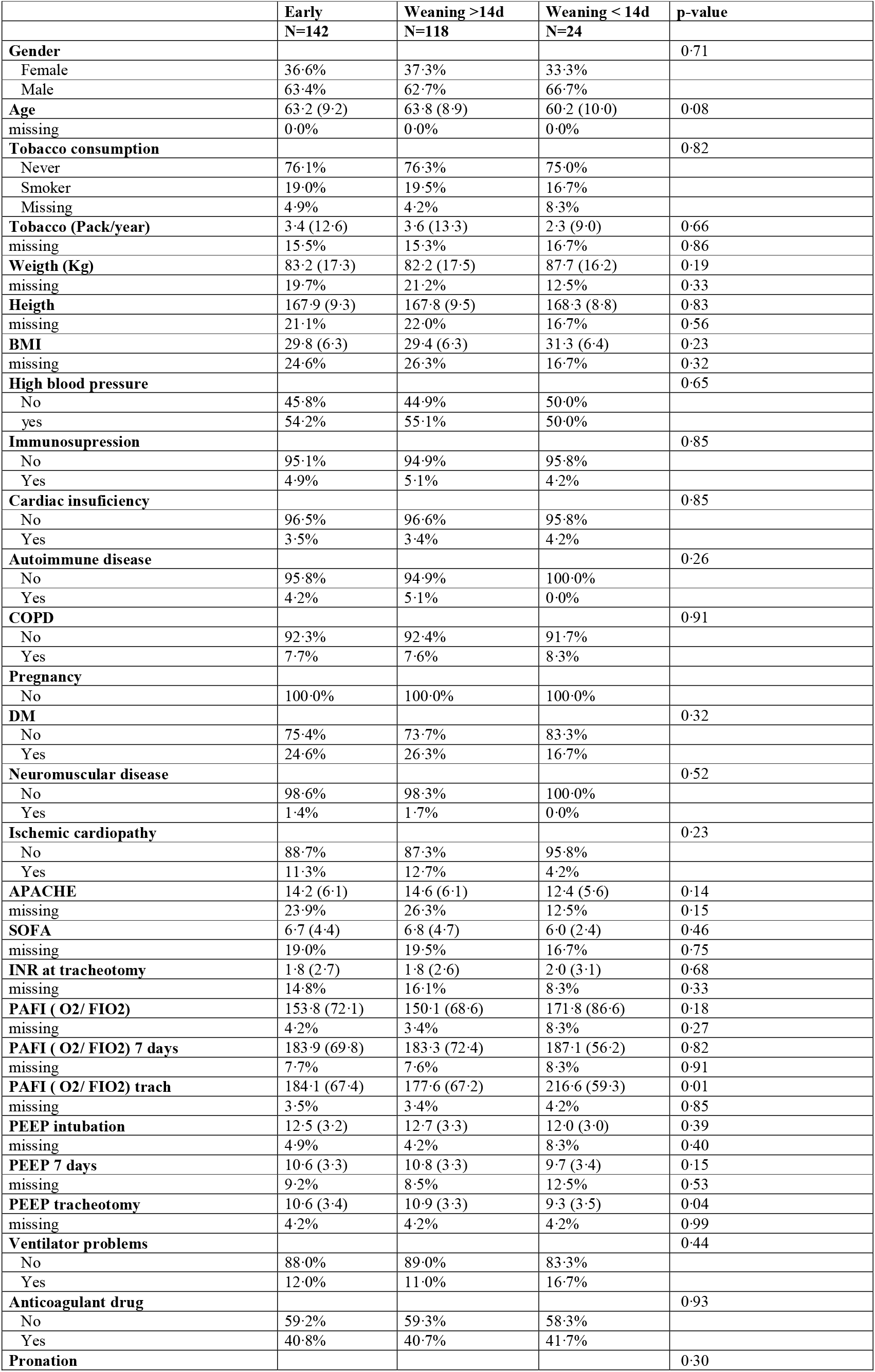
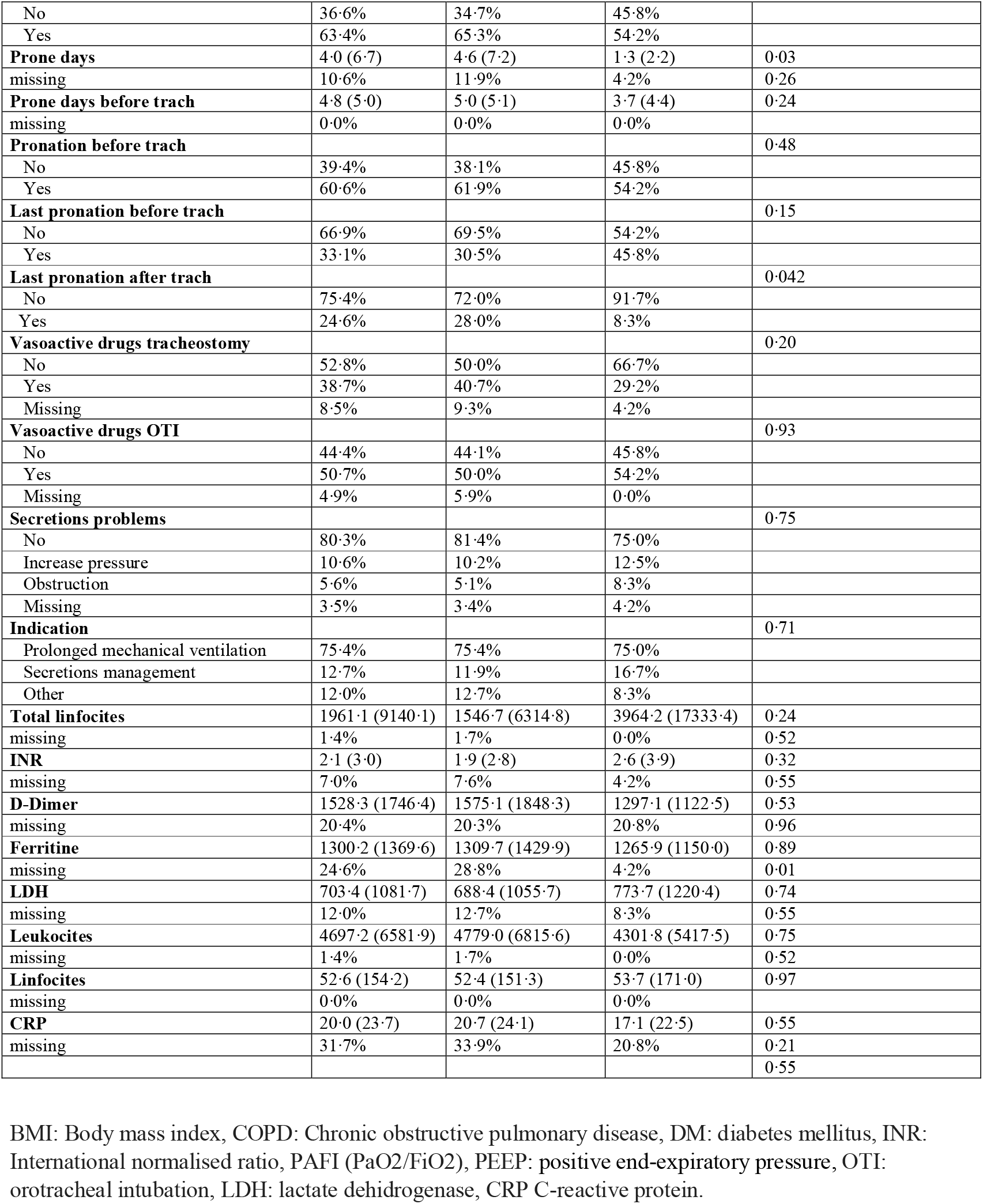
Baseline characteristics in early tracheotomised patients stratified by early weaning.

