## Supplementary Material for "Timing of elective tracheotomy and duration of mechanical ventilation amongst patients admitted to intensive care with severe COVID-19: a multicentre prospective cohort study"

**Table	S1. Baseline characteristics in tracheotomised patients stratified by exclusion**

|  | **Total** | **Included** | **General**  **Exclusions** | **p-value** | **Tracheostomy**  **<7 d** | **p-value** |
| --- | --- | --- | --- | --- | --- | --- |
|  | **N=794** | **N=754** | **N=40** |  | **N=58** |  |
| **Gender** |  |  |  | 0·16 |  | 0·32 |
| **Female** | 29·8% | 30·4% | 20·0% |  | 24·1% |  |
| **Male** | 70·2% | 69·6% | 80·0% |  | 75·9% |  |
| **Age (years)** | 62·9 (10·2) | 62·9 (10·2) | 62·8 (9·7) | 0·95 | 61·3 (11.1) | 0·22 |
| miss_age | 1·6% | 0·0% | 32·5% | <0·001 | 0·0% | 0·31 |
| **Tobacco consumption** |  |  |  | 0·12 |  | 0·35 |
| **Never** | 74·9% | 74·4% | 85·0% |  | 72·4% |  |
| **Smoker** | 16·0% | 16·4% | 7·5% |  | 20·7% |  |
| **Missing** | 9·1% | 9·2% | 7·5% |  | 6·9% |  |
| **Smoking Index** (pack/year) | 3·5 (12·8) | 3·5 (12·7) | 4·1 (15·3) | 0·77 | 4·9 (13·7) | 0·43 |
| Missing **_** | 16·0% | 16·4% | 7·5% | 0·13 | 12·1% | 0·40 |
| **Weigth** (Kg) | 83·1 (15·6) | 83·2 (15·6) | 81·4 (15·6) | 0·55 | 85·1 (17·0) | 0·37 |
| Missing | 20·0% | 19·6% | 27·5% | 0·23 | 19.0% | 0·83 |
| **Heigth** | 168·8 (9·2) | 168·8 (9·2) | 168·6 (9·4) | 0·90 | 17·.5 (10.5) | 0·044 |
| Missing | 22·2% | 21·9% | 27·5% | 0·40 | 22·4% | 0·96 |
| **BMI** | 29·3 (5·5) | 29·3 (5·5) | 28·8 (4·9) | 0·64 | 29·0 (5·9) | 0·71 |
| Missing | 24·2% | 23·7% | 32·5% | 0·21 | 24·1% | 0·99 |
| **HTA** |  |  |  | 0·43 |  | 0·64 |
| No | 53·8% | 53·4% | 60·0% |  | 56·9% |  |
| Yes | 45·8% | 46·2% | 40·0% |  | 43·1% |  |
| Missing | 0·4% | 0·4% | 0·0% |  | 0·0% |  |
| **Immunodepression** |  |  |  | 0·41 |  | 0·11 |
| No | 92·8% | 93·0% | 90·0% |  | 98·3% |  |
| Yes | 6·8% | 6·6% | 10·0% |  | 1·7% |  |
| Missing | 0·4% | 0·4% | 0·0% |  | 0·0% |  |
| **Cardiac insuficiency** |  |  |  | 0· 035 |  | 0·89 |
| No | 96·0% | 96·3% | 90·0% |  | 96·6% |  |
| Yes | 3·8% | 3·4% | 10·0% |  | 3·4% |  |
| Missing | 0·3% | 0·3% | 0·0% |  | 0·0% |  |
| **Autoimmune disease** |  |  |  | 0·11 |  | 0·32 |
| No | 94·1% | 93·8% | 100·0% |  | 91·4% |  |
| Yes | 5·7% | 6·0% | 0·0% |  | 8·6% |  |
| Missing | 0·3% | 0·3% | 0·0% |  | 0·0% |  |
| **COPD** |  |  |  | 0·58 |  | 0·67 |
| No | 9234% | 92·3% | 95·0% |  | 91·4% |  |
| Yes | 7·2% | 7·3% | 5·0% |  | 8·6% |  |
| Missing | 0·4% | 0·4% | 0·0% |  | 0·0% |  |
| **Pregnancy** |  |  |  | 0·068 |  | 0·57 |
| No | 99·4% | 99·5% | 97·5% |  | 100·0% |  |
| Yes | 0·5% | 0·4% | 2·5% |  | 0·0% |  |
| Missing | 0·1% | 0·1% | 0·0% |  | 0·0% |  |
| **Diabetes Mellitus** |  |  |  | 0·58 |  | 0·29 |
| No | 78·8% | 78·6% | 82·5% |  | 84·5% |  |
| Yes | 20·9% | 21·1% | 17·5% |  | 15·5% |  |
| Missing | 0·3% | 0·3% | 0·0% |  | 0·0% |  |
| **Neuromuscular disease** |  |  |  | 0·46 |  | 0·37 |
| No | 98·5% | 98·4% | 100·0% |  | 100·0% |  |
| Yes | 1·3% | 1·3% | 0·0% |  | 0·0% |  |
| Missing | 0·3% | 0·3% | 0·0% |  | 0·0% |  |
| **Ischemic cardiopathy** |  |  |  | 0·39 |  | <0·001 |
| No | 88·9% | 89·1% | 85·0% |  | 74·1% |  |
| Yes | 10·8% | 10·6% | 15·0% |  | 25·9% |  |
| Missing | 0·3% | 0·3% | 0·0% |  | 0·0% |  |
| **APACHE** | 15·0 (6·6) | 15·0 (6·6) | 15·1 (7·0) | 0·99 | 14·8 (6·9) | 0·81 |
| Missing | 18·8% | 19·5% | 5·0% | 0·022 | 34·5% | 0·001 |
| **SOFA** | 6·3 (3·8) | 6·3 (3·8) | 6·2 (3·3) | 0·93 | 8·4 (4·8) | <0·001 |
| Missing | 22·7% | 21·9% | 37·5% | 0·022 | 22·4% | 0·96 |
| **INR tracheotomy** | 1·5 (2·1) | 1·6 (2·1) | 1·4 (1·7) | 0·65 | 1·5 (2·0) | 0·98 |
| Missing | 16·8% | 17·0% | 12·5% | 0·46 | 6·9% | 0·037 |
| **PAFI ( O2/ FIO2)** | 140·6 (69·8) | 141·4 (69·9) | 123·5 (65·5) | 0·16 | 160·1 (77·6) | 0·036 |
| Missing | 11·2% | 10·7% | 20·0% | 0·071 | 10·3% | 0·83 |
| **PAFI ( O2/ FIO2) 7 days** | 183·8 (74·8) | 184·3 (74·8) | 173·3 (74·8) | 0·42 | 203·6 (88·3) | 0·068 |
| Missing | 14·4% | 14·1% | 20·0% | 0·30 | 24·1% | 0·027 |
| **PAFI ( O2/ FIO2) trach** | 193·7 (70·4) | 193·6 (70·2) | 194·4 (74·8) | 0·95 | 204·7 (80·3) | 0·23 |
| Missing | 7·8% | 7·4% | 15·0% | 0·082 | 5·2% | 0·44 |
| **PEEP intubation** | 12·6 (5·0) | 12·6 (5·0) | 12·7 (3·0) | 0·91 | 13·2 (3·8) | 0·39 |
| **miss_PEEPintub** | 11·0% | 10·6% | 17·5% | 0·17 | 8·6% | 0·55 |
| **PEEP 7 days** | 11·1 (7·4) | 11·1 (7·6) | 10·4 (3·8) | 0·60 | 11·3 (3·7) | 0·83 |
| miss_PEEP7 | 14·7% | 14·5% | 20·0% | 0·34 | 20·7% | 0·18 |
| **PEEP tracheotomy** | 9·8 (3·1) | 9·8 (3·1) | 9·7 (3·0) | 0·78 | 11·5 (3·9) | <0·001 |
| miss_PEEPtraq | 4·2% | 3·8% | 10·0% | 0·057 | 0·0% | 0·100 |
| **Ventilator problems** |  |  |  | 0·83 |  | 0·25 |
| No | 85·5% | 85·4% | 87·5% |  | 91·4% |  |
| Yes | 13·5% | 13·5% | 12·5% |  | 8·6% |  |
| Missing | 1·0% | 1·1% | 0·0% |  | 0·0% |  |
| **Anticoagulant drug** |  |  |  | 0·81 |  | 0·60 |
| No | 40·2% | 40·1% | 42·5% |  | 34·5% |  |
| Yes | 48·5% | 48·5% | 47·5% |  | 48·3% |  |
| Missing | 11·3% | 11·4% | 10·0% |  | 17·2% |  |
| **Pronation** |  |  |  | 0·11 |  | <0·001 |
| No | 31·5% | 32·1% | 20·0% |  | 60·3% |  |
| Yes | 68·5% | 67·9% | 80·0% |  | 39·7% |  |
| **Pronation days** | 5·8 (7·9) | 5·6 (7·7) | 8·3 (10·5) | 0·039 | 1·8 (3·6) | <0·001 |
| Missing | 10·1% | 10·5% | 2·5% | 0·10 | 5·2% | 0·20 |
| **Pronation days before trach** | 9·3 (9·2) | 9·1 (9·0) | 13·2 (12·2) | 0·005 | 1·6 (2·4) | <0·001 |
| Missing | 0·0% | 0·0% | 0·0% |  | 0·0% |  |
| **Pronation before_7d** |  |  |  | 0·063 |  | 0·005 |
| No | 36·3% | 37·0% | 22·5% |  | 53·4% |  |
| Yes | 63·7% | 63·0% | 77·5% |  | 46·6% |  |
| **Pronation days before 7d** | 3·9 (3·8) | 3·9 (3·8) | 4·2 (2·9) | 0·58 | 3·1 (4·1) | 0·076 |
| Missing | 0·3% | 0·0% | 5·0% | <0·001 | 0·0% | 0·69 |
| **Pronation before trach** |  |  |  | 0·47 |  | <0·001 |
| No | 35·3% | 35·5% | 30·0% |  | 67·2% |  |
| Yes | 64·7% | 64·5% | 70·0% |  | 32·8% |  |
| **Last pronation before tracheotomy** |  |  |  | 0·17 |  | <0·001 |
| No | 55·5% | 56·1% | 45·0% |  | 82·8% |  |
| Yes | 44·5% | 43·9% | 55·0% |  | 17·2% |  |
| **Last pronation after tracheotomy** |  |  |  | 0·63 |  | 0·91 |
| No | 80·5% | 80·6% | 77·5% |  | 81·0% |  |
| Yes | 19·5% | 19·4% | 22·5% |  | 19·0% |  |
| **Vasoactive drugs tracheostomy** |  |  |  | 0·30 |  | 0·55 |
| No | 47·4% | 46·8% | 57·5% |  | 46·6% |  |
| Yes | 40·6% | 40·8% | 35·0% |  | 46·6% |  |
| Missing | 12·1% | 12·3% | 7·5% |  | 6·9% |  |
| **Vasoactive drugs OTI** |  |  |  | 0·099 |  | 0·098 |
| No | 42·9% | 42·2% | 57·5% |  | 34·5% |  |
| Yes | 52·8% | 53·3% | 42·5% |  | 65·5% |  |
| Missing | 4·3% | 4·5% | 0·0% |  | 0·0% |  |
| **Secretions problems** |  |  |  | 0·82 |  | 0·37 |
| No | 74·4% | 74·4% | 75·0% |  | 82·8% |  |
| Increase pressure | 12·3% | 12·2% | 15·0% |  | 6·9% |  |
| Obstruction | 3·8% | 3·8% | 2·5% |  | 3·4% |  |
| Missing | 9·4% | 9·5% | 7·5% |  | 6·9% |  |
| **Indication tracheotomy** |  |  |  | 0·61 |  | 0·23 |
| Prolonged mechanical ventilation | 81·5% | 81·2% | 87·5% |  | 75·9% |  |
| Secretions management | 10·6% | 10·7% | 7·5% |  | 17·2% |  |
| Other | 7·8% | 8·0% | 5·0% |  | 6·9% |  |
| Missing | 0·1% | 0·1% | 0·0% |  | 0·0% |  |
| **Total linfocites** | 5238·0 (26329·0) | 5155·4 (26054·9) | 6808·5 (31439·1) | 0·70 | 3395·0 (12670·6) | 0·58 |
| missing | 1·8% | 1·7% | 2·5% | 0·72 | 0·0% | 0·29 |
| **INR** | 1·6 (2·2) | 1·6 (2·3) | 1·2 (0·2) | 0·21 | 1·6 (2·2) | 0·87 |
| missing | 7·8% | 8·1% | 2·5% | 0·20 | 3·4% | 0·20 |
| **D-Dimer** | 1499·8 (1708·0) | 1509·5 (1719·2) | 1299·6 (1468·7) | 0·53 | 1424·2 (1479·6) | 0·79 |
| missing | 23·8% | 23·5% | 30·0% | 0·35 | 39·7% | 0·003 |
| **Ferritine** | 1357·7 (1306·6) | 1345·5 (1292·6) | 1607·5 (1573·6) | 0·30 | 1114·0 (1043·2) | 0·17 |
| missing | 24·3% | 24·0% | 30·0% | 0·39 | 13·8% | 0·053 |
| **LDH** | 645·5 (837·1) | 651·0 (858·4) | 547·2 (221·3) | 0·47 | 1360·0 (1880·3) | <0·001 |
| missing | 14·4% | 14·6% | 10·0% | 0·42 | 24·1% | 0·027 |
| **Leukocites** | 4445·0 (9343·6) | 4448·9 (9494·0) | 4370·0 (5854·7) | 0·96 | 3396·3 (6162·1) | 0·37 |
| missing | 1·5% | 1·5% | 2·5% | 0·60 | 0·0% | 0·33 |
| **Linfocites** | 62·2 (183·5) | 61·2 (178·8) | 81·3 (259·0) | 0·51 | 52·2 (163·9) | 0·67 |
| missing | 1·4% | 1·3% | 2·5% | 0·54 | 0·0% | 0·35 |
| **CRP** | 20·0 (22·6) | 20·2 (22·7) | 17·3 (21·4) | 0·53 | 13·3 (21·1) | 0·044 |
| missing | 42·6% | 42·8% | 37·5% | 0·51 | 27·6% | 0·017 |

**Table	S2. Pronation and pronation days before tracheotomy in tracheostomised patients stratified by early or late weaning.**

|  | **Total** | **Late**  **(>10d after IOT)** | **Early**  **(10d after IOT)** | **p-value** |
| --- | --- | --- | --- | --- |
|  | N=696 | N=554 | N=142 |  |
| **Pronation anytime** | 70·3% | 72·0% | 63·4% | 0·04 |
| **Days of pronation** | 6·0 (7·8) | 6·5 (8·0) | 4·0 (6·7) | 0·00 |
| **Missing** | 10·9% | 11·0% | 10·6% | 0·88 |
| **Pronation on 7 days post-OTI** | 64·4% | 65·2% | 61·3% | 0·39 |
| **Pronation days on 7 days post-OTI** | 4·0 (3·8) | 4·0 (3·7) | 3·8 (4·1) | 0·64 |
| **Pronation before tracheotomy** | 67·1% | 68·8% | 60·6% | 0·06 |
| **Days of pronation before tracheotomy** | 9·7 (9·1) | 11·0 (9·4) | 4·8 (5·0) | <0·001 |
| **Last pronation before tracheotomy** | 46·1% | 49·5% | 33·1% | <0·001 |
| **Pronation continued after tracheotomy** | 19·4% | 18·1% | 24·6% | 0·08 |

**Figure S3. Additional Analyses of time to weaning: Fine and Gray competing risks model and 14-day Poisson**

|  | **sdHR** | **CI95** |  |  | **CI95** |  |
| --- | --- | --- | --- | --- | --- | --- |
| *Fine and Gray Crude* | 1·22 | (0·98 | 1·52) |  |  |  |
| *F-G Age and Gender Adjusted* | 1·21 | (0·97 | 1·50) |  |  |  |
| *F-G Fully Adjusted* | 1·16 | (0·92 | 1·47) |  |  |  |
|  | **RR** |  |  | **RR** |  |  |
| *14-day Poisson* | 1·21 | (0·92 | 1·60) | 0·93 | (0·58 | 1·50) |

**Figure S4. Baseline characteristics in early tracheotomised patients stratified by early weaning**

|  | **Early** | **Weaning >14d** | **Weaning < 14d** | **p-value** |
| --- | --- | --- | --- | --- |
|  | **N=142** | **N=118** | **N=24** |  |
| **Gender** |  |  |  | 0·71 |
| Female | 36·6% | 37·3% | 33·3% |  |
| Male | 63·4% | 62·7% | 66·7% |  |
| **Age** | 63·2 (9·2) | 63·8 (8·9) | 60·2 (10·0) | 0·08 |
| missing | 0·0% | 0·0% | 0·0% |  |
| **Tobacco consumption** |  |  |  | 0·82 |
| Never | 76·1% | 76·3% | 75·0% |  |
| Smoker | 19·0% | 19·5% | 16·7% |  |
| Missing | 4·9% | 4·2% | 8·3% |  |
| **Tobacco (Pack/year)** | 3·4 (12·6) | 3·6 (13·3) | 2·3 (9·0) | 0·66 |
| missing | 15·5% | 15·3% | 16·7% | 0·86 |
| **Weigth (Kg)** | 83·2 (17·3) | 82·2 (17·5) | 87·7 (16·2) | 0·19 |
| missing | 19·7% | 21·2% | 12·5% | 0·33 |
| **Heigth** | 167·9 (9·3) | 167·8 (9·5) | 168·3 (8·8) | 0·83 |
| missing | 21·1% | 22·0% | 16·7% | 0·56 |
| **BMI** | 29·8 (6·3) | 29·4 (6·3) | 31·3 (6·4) | 0·23 |
| missing | 24·6% | 26·3% | 16·7% | 0·32 |
| **High blood pressure** |  |  |  | 0·65 |
| No | 45·8% | 44·9% | 50·0% |  |
| yes | 54·2% | 55·1% | 50·0% |  |
| **Immunosupression** |  |  |  | 0·85 |
| No | 95·1% | 94·9% | 95·8% |  |
| Yes | 4·9% | 5·1% | 4·2% |  |
| **Cardiac insuficiency** |  |  |  | 0·85 |
| No | 96·5% | 96·6% | 95·8% |  |
| Yes | 3·5% | 3·4% | 4·2% |  |
| **Autoimmune disease** |  |  |  | 0·26 |
| No | 95·8% | 94·9% | 100·0% |  |
| Yes | 4·2% | 5·1% | 0·0% |  |
| **COPD** |  |  |  | 0·91 |
| No | 92·3% | 92·4% | 91·7% |  |
| Yes | 7·7% | 7·6% | 8·3% |  |
| **Pregnancy** |  |  |  |  |
| No | 100·0% | 100·0% | 100·0% |  |
| **DM** |  |  |  | 0·32 |
| No | 75·4% | 73·7% | 83·3% |  |
| Yes | 24·6% | 26·3% | 16·7% |  |
| **Neuromuscular disease** |  |  |  | 0·52 |
| No | 98·6% | 98·3% | 100·0% |  |
| Yes | 1·4% | 1·7% | 0·0% |  |
| **Ischemic cardiopathy** |  |  |  | 0·23 |
| No | 88·7% | 87·3% | 95·8% |  |
| Yes | 11·3% | 12·7% | 4·2% |  |
| **APACHE** | 14·2 (6·1) | 14·6 (6·1) | 12·4 (5·6) | 0·14 |
| missing | 23·9% | 26·3% | 12·5% | 0·15 |
| **SOFA** | 6·7 (4·4) | 6·8 (4·7) | 6·0 (2·4) | 0·46 |
| missing | 19·0% | 19·5% | 16·7% | 0·75 |
| **INR de traqueo** | 1·8 (2·7) | 1·8 (2·6) | 2·0 (3·1) | 0·68 |
| missing | 14·8% | 16·1% | 8·3% | 0·33 |
| **PAFI ( O2/ FIO2)** | 153·8 (72·1) | 150·1 (68·6) | 171·8 (86·6) | 0·18 |
| missing | 4·2% | 3·4% | 8·3% | 0·27 |
| **PAFI ( O2/ FIO2) 7 days** | 183·9 (69·8) | 183·3 (72·4) | 187·1 (56·2) | 0·82 |
| missing | 7·7% | 7·6% | 8·3% | 0·91 |
| **PAFI ( O2/ FIO2) trach** | 184·1 (67·4) | 177·6 (67·2) | 216·6 (59·3) | 0·01 |
| missing | 3·5% | 3·4% | 4·2% | 0·85 |
| **PEEP intubation** | 12·5 (3·2) | 12·7 (3·3) | 12·0 (3·0) | 0·39 |
| missing | 4·9% | 4·2% | 8·3% | 0·40 |
| **PEEP 7 days** | 10·6 (3·3) | 10·8 (3·3) | 9·7 (3·4) | 0·15 |
| missing | 9·2% | 8·5% | 12·5% | 0·53 |
| **PEEP tracheostomy** | 10·6 (3·4) | 10·9 (3·3) | 9·3 (3·5) | 0·04 |
| missing | 4·2% | 4·2% | 4·2% | 0·99 |
| **Ventilator problems** |  |  |  | 0·44 |
| No | 88·0% | 89·0% | 83·3% |  |
| Yes | 12·0% | 11·0% | 16·7% |  |
| **Anticoagulant drug** |  |  |  | 0·93 |
| No | 59·2% | 59·3% | 58·3% |  |
| Yes | 40·8% | 40·7% | 41·7% |  |
| **Pronation** |  |  |  | 0·30 |
| No | 36·6% | 34·7% | 45·8% |  |
| Yes | 63·4% | 65·3% | 54·2% |  |
| **Prone days** | 4·0 (6·7) | 4·6 (7·2) | 1·3 (2·2) | 0·03 |
| missing | 10·6% | 11·9% | 4·2% | 0·26 |
| **Prone days before trach** | 4·8 (5·0) | 5·0 (5·1) | 3·7 (4·4) | 0·24 |
| missing | 0·0% | 0·0% | 0·0% |  |
| **Pronation before trach** |  |  |  | 0·48 |
| No | 39·4% | 38·1% | 45·8% |  |
| Yes | 60·6% | 61·9% | 54·2% |  |
| **Last pronation before trach** |  |  |  | 0·15 |
| No | 66·9% | 69·5% | 54·2% |  |
| Yes | 33·1% | 30·5% | 45·8% |  |
| **Last pronation after trach** |  |  |  | 0·042 |
| No | 75·4% | 72·0% | 91·7% |  |
| Yes | 24·6% | 28·0% | 8·3% |  |
| **Vasoactive drugs tracheostomy** |  |  |  | 0·20 |
| No | 52·8% | 50·0% | 66·7% |  |
| Yes | 38·7% | 40·7% | 29·2% |  |
| Missing | 8·5% | 9·3% | 4·2% |  |
| **Vasoactive drugs OTI** |  |  |  | 0·93 |
| No | 44·4% | 44·1% | 45·8% |  |
| Yes | 50·7% | 50·0% | 54·2% |  |
| Missing | 4·9% | 5·9% | 0·0% |  |
| **Secretions problems** |  |  |  | 0·75 |
| No | 80·3% | 81·4% | 75·0% |  |
| Increase pressure | 10·6% | 10·2% | 12·5% |  |
| Obstruction | 5·6% | 5·1% | 8·3% |  |
| Missing | 3·5% | 3·4% | 4·2% |  |
| **Indication** |  |  |  | 0·71 |
| Prolonged mechanical ventilation | 75·4% | 75·4% | 75·0% |  |
| Secretions management | 12·7% | 11·9% | 16·7% |  |
| Other | 12·0% | 12·7% | 8·3% |  |
| **Total linfocites** | 1961·1 (9140·1) | 1546·7 (6314·8) | 3964·2 (17333·4) | 0·24 |
| missing | 1·4% | 1·7% | 0·0% | 0·52 |
| **INR** | 2·1 (3·0) | 1·9 (2·8) | 2·6 (3·9) | 0·32 |
| missing | 7·0% | 7·6% | 4·2% | 0·55 |
| **D-Dimer** | 1528·3 (1746·4) | 1575·1 (1848·3) | 1297·1 (1122·5) | 0·53 |
| missing | 20·4% | 20·3% | 20·8% | 0·96 |
| **Ferritine** | 1300·2 (1369·6) | 1309·7 (1429·9) | 1265·9 (1150·0) | 0·89 |
| missing | 24·6% | 28·8% | 4·2% | 0·01 |
| **LDH** | 703·4 (1081·7) | 688·4 (1055·7) | 773·7 (1220·4) | 0·74 |
| missing | 12·0% | 12·7% | 8·3% | 0·55 |
| **Leukocites** | 4697·2 (6581·9) | 4779·0 (6815·6) | 4301·8 (5417·5) | 0·75 |
| missing | 1·4% | 1·7% | 0·0% | 0·52 |
| **Linfocites** | 52·6 (154·2) | 52·4 (151·3) | 53·7 (171·0) | 0·97 |
| missing | 0·0% | 0·0% | 0·0% |  |
| **CRP** | 20·0 (23·7) | 20·7 (24·1) | 17·1 (22·5) | 0·55 |
| missing | 31·7% | 33·9% | 20·8% | 0·21 |
|  |  |  |  | 0·55 |
